# A novel cohort analysis approach to determining the case fatality rate of COVID-19 and other infectious diseases

**DOI:** 10.1101/2020.04.02.20051482

**Authors:** Charit Samyak Narayanan

**Author notes:** These authors contributed equally to this work. Mission San Jose High School, 41717 Palm Ave, Fremont, California, United States of America.

## Abstract

As the Coronavirus contagion develops, it is increasingly important to understand the dynamics of the disease. Its severity is best described by two parameters: its ability to spread and its lethality. Here, we combine a mathematical model with a cohort analysis approach to determine the range of case fatality rates (CFR). We use a logistical function to describe the exponential growth and subsequent flattening of COVID-19 CFR that depends on three parameters: the final CFR (L), the CFR growth rate (k), and the onset-to-death interval (t_0_). Using the logistic model with specific parameters (L, k and t_0_), we calculate the number of deaths each day for each cohort. We build an objective function that minimizes the root mean square error between the actual and predicted values of cumulative deaths and run multiple simulations by altering the three parameters. Using all of these values, we find out which set of parameters returns the lowest error when compared to the number of actual deaths. We were able to predict the CFR much closer to reality at all stages of the viral outbreak compared to traditional methods. This model can be used far more effectively than current models to estimate the CFR during an outbreak, allowing for better planning. The model can also help us better understand the impact of individual interventions on the CFR. With much better data collection and labeling, we should be able to improve our predictive power even further.

## Introduction

The Coronavirus Disease 2019, or COVID-19, is a new virus that causes respiratory ailments [1]. Its origins can most likely be traced back to Wuhan, China, where it was first reported in December of 2019. It is comparable to other zoonotic coronaviruses such as SARS-CoV and MERS-CoV [2]. Although most earlier cases had been confined to mainland China, notably Hubei Province, a considerable number of people have been infected all over the world since, and the WHO characterized the outbreak as a pandemic on March 11 [1]. As of March 17, over 179,000 have been confirmed to be infected, with almost 7400 deaths [3].

As the Coronavirus contagion develops, with its rapid spread inciting concerns over the epidemic’s global impact, it is increasingly important to understand the dynamics of the disease. Its severity is best described by two concepts: its ability to spread and its lethality. Its transmissibility is best measured by R0 (the basic reproduction number), the average number of people to whom an infected person spreads the disease to [4]. Its Case Fatality Rate (CFR) reveals the proportion of people who die from disease among all individuals who contract the disease [5].

Knowing the CFR is essential because it is the single most accurate metric to appraise the threat posed by infectious diseases such as COVID-19 and predict its prognosis. Although R0 is important to understand the rate of spread and contagiousness, it does not describe the lethality of a disease. The case incidence would describe the prevalence of the disease, but if it has a negligible CFR, the severity of the disease is likely low. During the 2015 MERS outbreak in South Korea, there were only 186 cases, but the number of fatalities that resulted (38) was an adequate cause for alarm [6]. Thus, the CFR, if calculated correctly, can help inform the appropriate measures to control the epidemic.

The inaccuracies related to calculating the CFR during an ongoing disease outbreak are a known problem. There are primarily three methods that have been discussed in literature pertaining to the evaluation of the CFR. A simple statistical estimate is evaluated by dividing the aggregate number of deaths over the aggregate confirmed cases. This is problematic because it ignores the lag effect; people become infected on different days and the mix of their onset-to-death intervals is not taken into account. The second method calculates the CFR as the ratio of aggregate deaths to closed cases. This ignores those who are still sick and only includes those who have had a definite outcome (death or recovery). The third procedure [5] uses the Kaplan-Meier survival procedure, jointly considering two the probability of two outcomes (death and recovery), which is violated if mean duration from hospital admission to death is substantially shorter than the mean duration from hospital admission to discharge. Here, we develop a novel procedure that evaluates the risk of death among confirmed cases, explicitly accounting for the time of the onset-to-death interval. This method, in principle, should yield the best results. All these methods except the Kaplan-Meier survival procedure will without converge in the future when the outbreak is no longer in-progress but diverge during the outbreak.

To execute our proposed cohort-based method for calculating the CFR during an ongoing outbreak, we explicitly model the death rates using a growth function to simulate multiple conditions. This is done by specifically altering time from illness to death and the slope of the growth of deaths. Our approach has multiple advantages over the other methods. We can identify and compare the CFR of each cohort as well as its evolution over time. Also, we can calculate the CFR earlier in the stage of disease development, especially if we know the time from infection to time of death. Our method can help increase the accuracy of calculated CFR for both COVID-19 and future viral diseases. This analysis will assist in furthering efforts to halt the spread of COVID-19 and drive an informed, comprehensive approach towards counteracting its spread. The range of CFR possibilities and onset-to-death intervals will also raise awareness among the public so that the correct measures are taken.

## Methods

### Data sources

We used publicly available datasets [7] on cases in Hubei province, China. Specifically, we used the number of confirmed cases, new cases, recovery cases, and deaths from January 22, 2020 to March 11, 2020.

### Cohort-based Method

Cohort analysis has been used in improving product retention [8] for many years. In this piece, we apply this approach to describe the CFR over time for individual cohorts. This would assist in keeping track of the CFR’s evolution and the impact of interventions and other preventative/reactive measures.

A simulated Triangle chart best illustrates cohort CFRs. In Fig 1, the uppermost rows depict older cohorts, and newer cohorts are represented by the lower rows. Each column represents an additional day after they were infected (moving from left to right), with the number in the box representing the cumulative CFR (cumulative deaths over the number of people infected) within the cohort up to the day. Scanning each row from left to right gives each individual cohort’s most recent CFR. Scanning down each column yields the CFR for each cohort for exactly the same number of days of the development of the infection. This can tell us about the effects of interventions if the CFR abruptly rises or drops. For example, when Wuhan, China was placed under lockdown, the CFR would have decreased in the following days. Scanning diagonally reveals each cohort’s CFR on the same day. The intervention’s effects should also manifest on the diagonal. Thus, with the right level of information, we would be able to identify if the intervention measures are effective.

**Fig 1.**
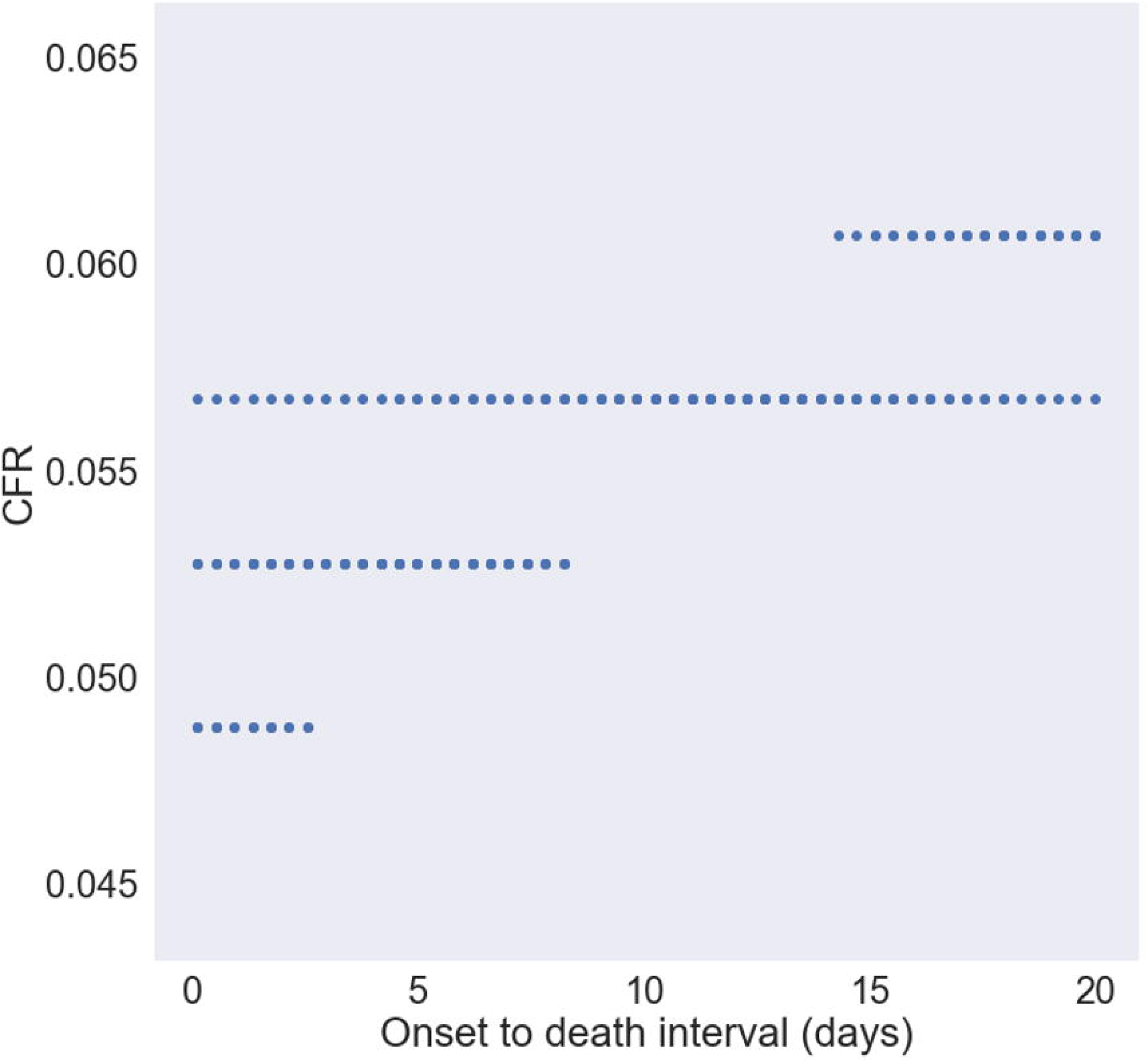
Simulated triangle chart that illustrates the cohort method. Each cell contains the CFR for a given cohort on a given day. Each row represents a cohort who experienced the onset of disease on the same day.

If the groups are homogenous and identical to each other in every way (have the same mix), the cohorts would have the same CFR. This is because each cohort CFR would change at the same rate, assuming they are the same age, have the similar comorbidities (if any), etc. However, in reality this will likely not be the case since groups will naturally have people with differing characteristics and their mix will affect the CFR. In an idealistic sense, we would be able to have homogenous cohorts, filtered by certain conditions (age group, smoking habits, etc.) and looking at the cohorts over time, we would be able to establish a pattern, and obtain a reliable CFR [ [9], [10]].

We would need to construct these triangle charts for groups that are as homogeneous as possible to ensure that certain members do not skew the death rates. Age, high-risk medical groups, and countries are the right granular segments to look at. Using these, we can most accurately gauge the case fatality rate for each group.

### Case Fatality Rate Model

The case fatality rate as a function of time since infection can be modeled in multiple different ways [11]. We use a logistical function to describe the exponential growth, and subsequent flattening, of COVID-19 case fatality rate; it is the simplest form that satisfies the S-shaped growth [12]. This can be expressed as:

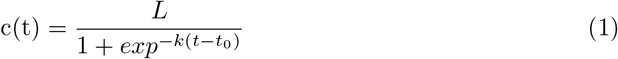

In this case, c(t) represents the CFR as a function of days since infection and depends on three parameters: the final CFR (L), the CFR growth rate (k), and the onset-to-death interval (t_0_). The final CFR is the case fatality rate when the epidemic has run its course; a larger CFR would imply that a higher proportion of people eventually die. The CFR growth rate is the slope of case fatality rate; a greater slope reveals a quicker progression to death. t_0_ is the inflection point and indicative of the onset-to-death interval; a greater t_0_ reveals a longer progression to death.

Using the logistic model with specific parameters, we calculate the number of deaths each day for each cohort by multiplying the case fatality rate by the number of cases (or cohort size). We then create a triangle chart of the number of deaths for each cohort. By adding along the diagonal, we are able to estimate the number of deaths from all cohorts on any given day. Thus, given a set of values from L, K and t_0_ we are able to estimate the aggregate number of deaths from all cohorts.

Finally, we build an objective function that minimizes the root mean square error between the actual and predicted values of cumulative deaths and run multiple simulations by altering the three parameters. Using all of these values, we find out which set of parameters returns the lowest error when compared to the number of actual deaths. Using this methodology, we determine the probable range of case fatality rates.

## Results

### Best fit

We ran 125,000 simulations, modifying the onset-to-death interval, the CFR, and the CFR growth rate. The CFR was kept in the range of 0.5% to 20.0%; the slope was held between 0·005 and 0·20; and the onset-to-death interval was simulated between 0.1 and 20·0. We picked these ranges after some modeling exploration that convinced us that the solution would certainly be within these parameters. We calculated the coefficient of determination (R_2_) for all of these simulations and identified the value for which the fit was the best. We show results from two cases: one where we are relatively early in the disease phase and another that is later. We studied these two cases to understand how the progression of the outbreak affected the results.

Fig 2 compares the modeled and actual number of cumulative deaths for the best fit for the late stage case. For this simulation, we used data from Jan 22 through March 11. The model is able to fit the actual data extremely well with a very high R_2_ of 0·998, indicating that the model and parameters chosen are strongly representative of the reality. Specifically, the model closely matches the actual data where the CFR is 4·88%, the number of days to the point of inflection is 3·35, and the slope is 0·088.

**Fig 2.**
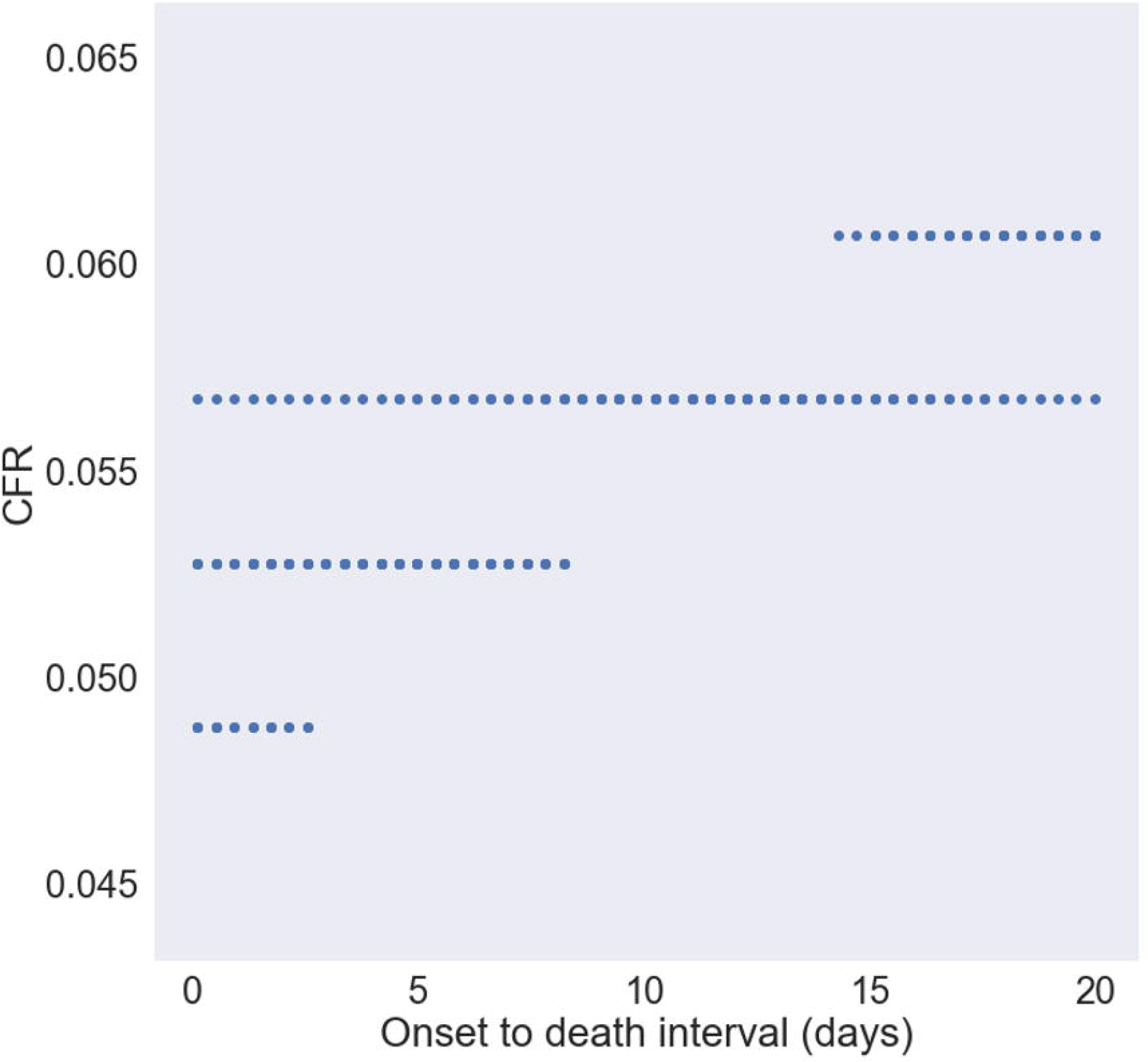
Comparison of modeled and actual number of deaths for the late stage case. Used data from January 22 through March 11

Fig 3 compares the modeled and actual number of cumulative deaths for the best fit for the early stage case. For this simulation, we used data from January 22 through February 8. Again, the model is able to fit the actual data extremely well with a very high R_2_ of 0·986. In this case, CFR is 5·67 %, the number of days to the point of inflection is 10·3, and the slope is 0·01.

**Fig 3.**
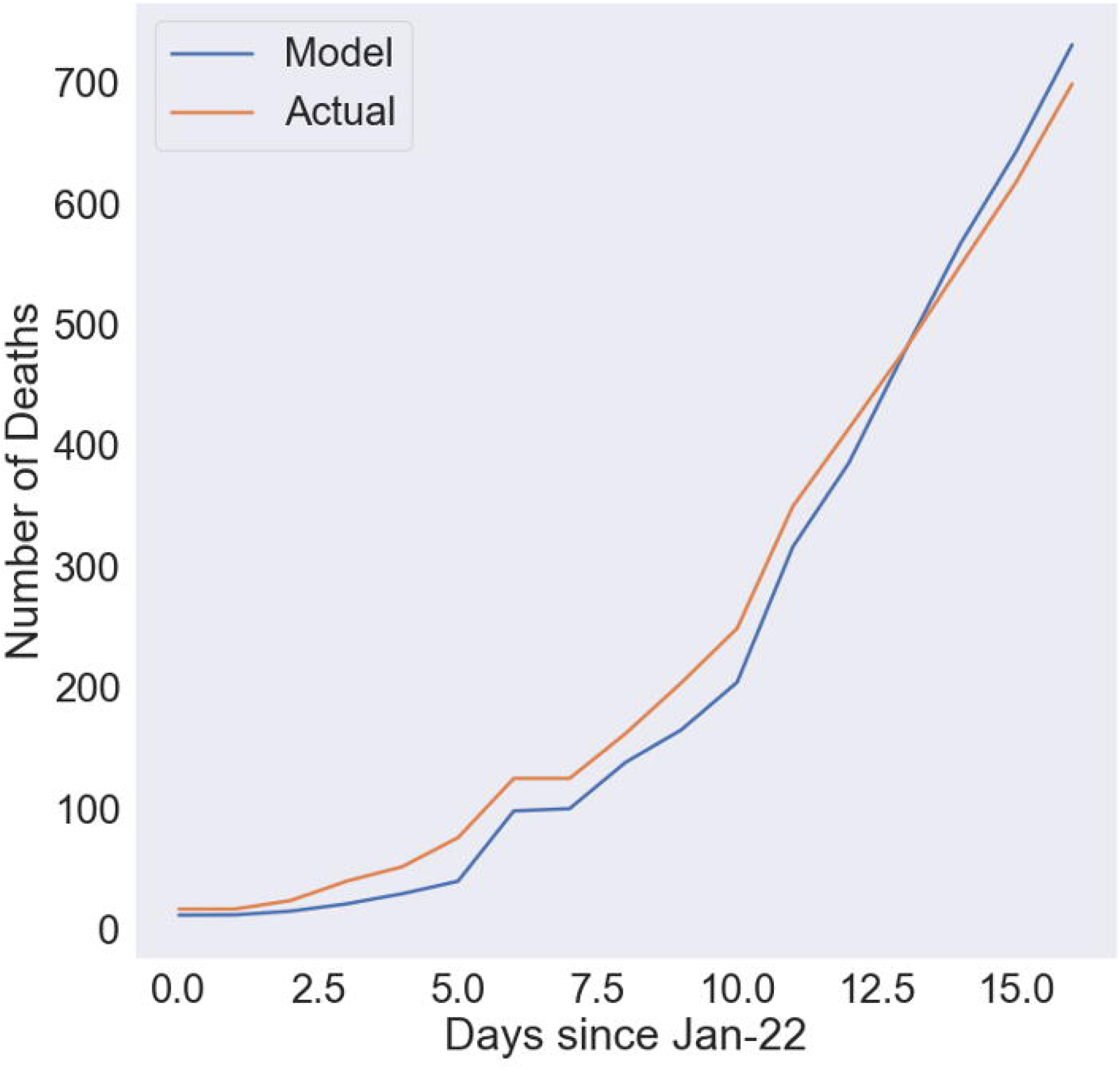
Comparison of modeled and actual number of deaths for the early stage case. Used data from January 22 through February 8.

The difference in decrease in case fatality rates could be due to a number of factors including the tactics employed by the Chinese government. These two cases demonstrate the strong predictive power of the cohort-based methodology. Thus, using a combination of known cohort sizes and the number of deaths on each day, we are able to predict mortality very well.

### Simulations

Fig 4 shows combinations of onset-to-death interval and case fatality rate under which the R_2_ between the model and reality is ¿= 0·98 for the shorter time frame (up to Feb 8). This demonstrates that the range of possibilities for the case fatality rate is from 4·8% to 6·1%. Furthermore, the case fatality rate is most likely between 5·5% and 6% due to its higher frequency. There is a wide range of values of onset to death that are theoretically possible. However, if we know the onset to death to be 10%, then the only possible case fatality rate that satisfies this is a case fatality rate of 5·7%. Therefore, a very important implication of this work is that we can accurately ascertain the case fatality rate given the onset-to-death interval. The practical implication is to calculate this value accurately and evaluate then ascertain case fatality rate.

**Fig 4.**
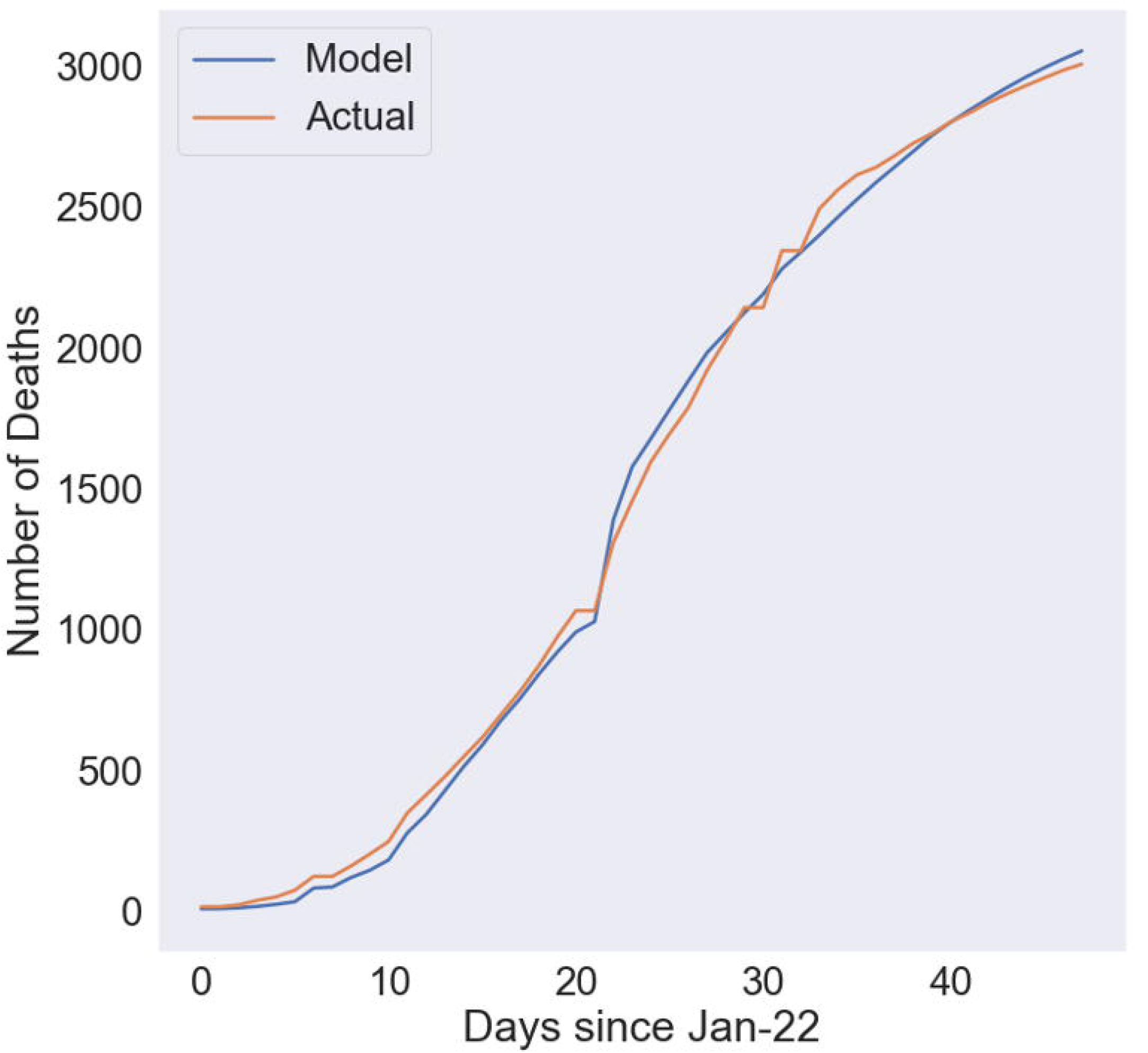
Onset-to-death interval and case fatality rate that yield the best fit. The parameters where R_2_ ¿= 0·98 for the early stage case. Demonstrates that the range of possibilities for the case fatality rate is from 4·8% to 6·1%.

### Comparison of Methods

We compare three methods (Fig 5): the completed case fatality rate (deaths/ (deaths + recovered)), the simple case fatality rate (deaths/confirmed), and the cohort-based mortality defined in this work. The completed case fatality rate ignores people in the midst of the infection and the simple method ignores the lag effect. The completed case fatality rate is very high at the start of an outbreak as very few have recovered compared to the amount of people who died. However, as the outbreak spreads, the number of recoveries begins to outpace the number of deaths. As the number of outstanding cases begin to reduce toward the end of an outbreak, this method converges to the true case fatality rate.

**Fig 5.**
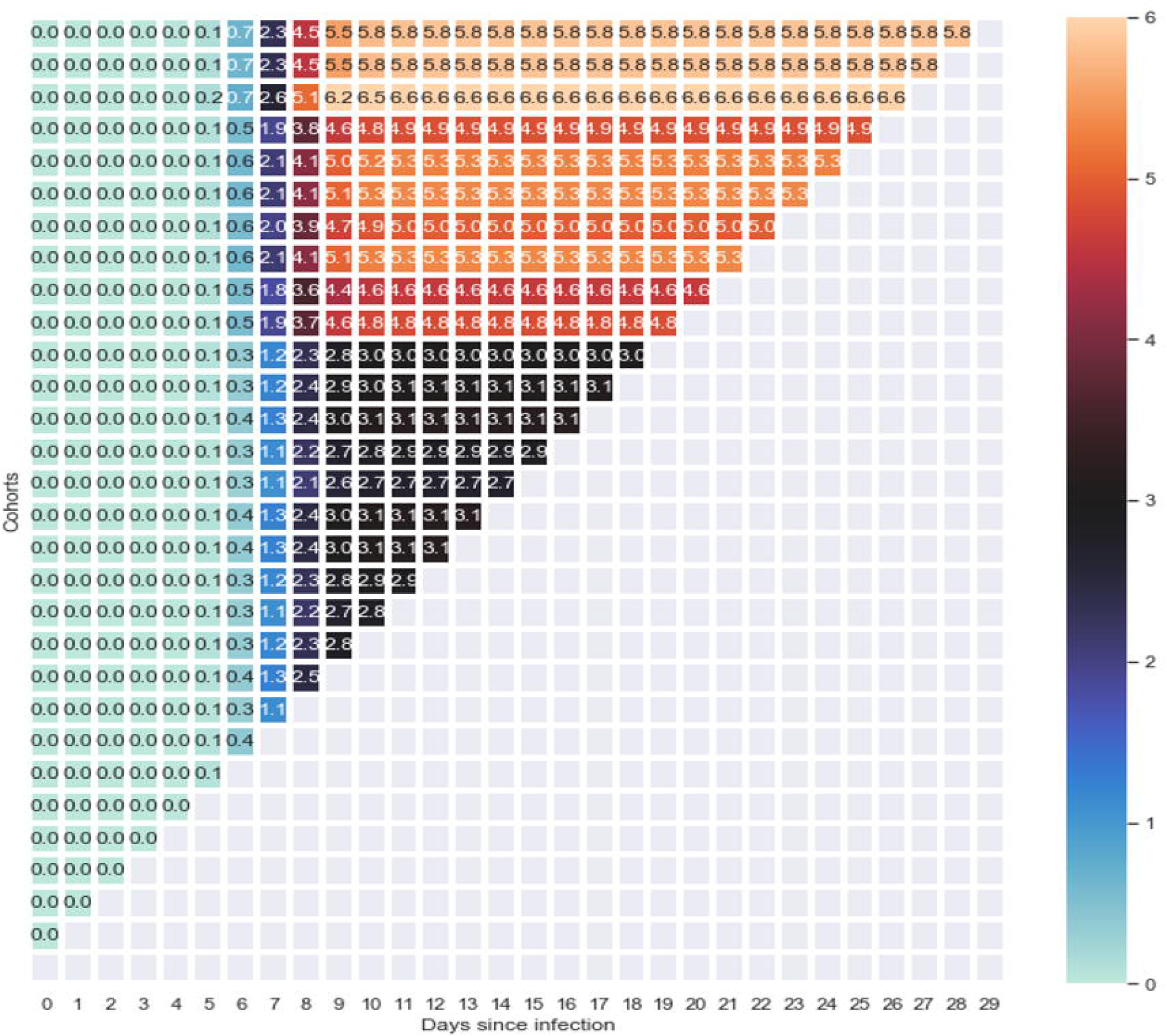
Comparison of CFR calculated with 3 methods. The three methods: completed CFR (deaths/ (deaths + recovered)); the simple CFR (deaths/confirmed); the cohort-based CFR defined in this work. Onset-to-death interval and case fatality rate that yield the best fit.

The simple case fatality rate begins low primarily because there are likely many more confirmed cases than deaths. Over time, as the deaths begin to increase, this ratio increases as well. As the number of new cases start to diminish, this ratio begins to converge to the true case fatality rate.

Although they are both erroneous in the midst of an outbreak, they are accurate when the outbreak is over: when everyone who has ever contracted it either recovers or dies, and no one new contracts it. In this situation, the CFR calculated with the simple method would equal the CFR calculated with the completed method. This is because the only difference between the two is that the second method includes ongoing cases. So, when the epidemic is over, there will be no ongoing cases and they will yield the same CFR.

The best-fit cohort-based method produces a case fatality rate that lies between the simple and completed methods. It produces far more realistic estimates at the earlier stages than the other two methods. Additionally, towards the later stages, it begins to converge much more quickly. Moreover, it appears that the other two methods are beginning to converge towards the cohort-based method.

## Conclusion

We combined a mathematical model with a cohort analysis approach to determine the range of CFR. We were able to predict the CFR much closer to reality at all stages of the viral outbreak compared to traditional methods such as dividing deaths by all closed cases and dividing deaths by the total number of cases. We also estimated that the mortality in Hubei province declined with the passage of time very likely due to government interventions.

Our analyses highlight the importance of collecting data such as the onset-to-death interval which will help us narrow the range of possibilities of the CFR early in the outbreak. Furthermore, using the triangle chart methodology and obtaining additional granular data, we are able to understand the evolution of the outbreak and the effect of interventions on the CFR. Moreover, even without the granular information of the progression of the disease (such as cough or fever) at the individual level, we are able to predict the CFR quite well.

However, numerous factors pose challenges to calculating the CFR using the cohort method. Firstly, the date of infection is difficult to determine since the onset-to-death interval is still uncertain. This is because there are numerous stages to contracting a disease: initially contracting the disease, showing symptoms, reporting the symptoms, being tested, getting results, going to a hospital and reaching an outcome. If different agencies measure the onset differently, there can be high degrees of bias. Defining a common terminology and language and measuring at all the stages is very important for future progress in estimating mortality.

Additionally, the CFR varies significantly with factors such as age, smoking habits, etc. If the proportions of differing-risk groups in each infected cohort varies, the CFR will be mis-representative [ [9], [10]]

Finally, the risk of mortality varies by place. With any rapidly spreading contagion, the samples chosen for determining the CFR are biased since the virus propagates so quickly, affecting different people in different places. So far, the CFR is heavily skewed by the numbers from Hubei Province, where the majority of cases originate. As the virus moves to different places, the mortality will inherently be different, and must, therefore, be representative of the threat posed so adequate precautions can be taken. For example, the danger it poses to people in developing countries would be significantly higher than the danger to people in developed countries, due to factors such as the people’s immunity/health, sanitation, access to medical services, etc. Thus, to give an accurate appraisal of the threat, there should be cohorts organized by place as well.

Even with all the above limitations, the cohort-based methodology is much better in estimating the case fatality rates and should be used for the current outbreak of COVID-19 in other regions as well as for future outbreaks. We should additionally modify the cohort-based methodology to model recovered patients in addition to the dead which would help us reach higher degrees of accuracy in calculating CFR.

## Data Availability

All data are fully available without restriction. All data used is from the database listed below.

https://github.com/CSSEGISandData/COVID-19

